# Pilot Study of Smartphone Ecological Momentary Assessment and Wearable Activity Tracking in Pediatric Depression

**DOI:** 10.1101/2025.02.10.25321998

**Authors:** Jimena Unzueta Saavedra, Emma A. Deaso, Margot Austin, Laura Cadavid, Rachel Kraff, Emma. E. M. Knowles

## Abstract

**Background:** Adolescent depression is a significant public health concern. The presentation of depressive symptoms varies widely among individuals, fluctuating in intensity over time. EMA offers a unique advantage by enhancing ecological validity and reducing recall bias, allowing for a more accurate and nuanced understanding of MDD symptoms. This methodology provides valuable insights into the fluctuating nature of depression, which could inform more personalized and timely interventions.

**Objectives:** This study aims to: (1) evaluate the feasibility of collecting smartphone-based Ecological Momentary Assessment (EMA) data alongside activity and sleep tracking in adolescents with depression; (2) investigate the severity and variability of mood symptoms reported over time; and (3) explore the relationship between mood, activity, and sleep.

**Methods:** Thirty-six participants (23 with Major Depressive Disorder (MDD), 13 unaffected controls; 75% female, mean age 19.50 years) completed twice-daily EMA check-ins over two weeks, complemented by continuous activity and sleep monitoring using FitBit Charge 3 devices. The study examined feasibility, usability of EMA app, symptom severity and variability, and relationships between mood, activity, and sleep. We applied linear mixed-effects regression to the data to examine relationships between variables.

**Results:** Participants completed a total of 923 unique checkins (mean check-ins per participant = 25.60). Overall compliance rates were high (91.57%) indicating the approach is highly feasible. MDD participants demonstrated greater symptom severity and variability over time compared to controls (*β* = 34.48, *p* = 2.17×10^−06^). Individuals with MDD exhibited greater diurnal variation (*β* = −2.54, p = 5.14×10^−03^) with worse mood in the morning and worse mood than anxiety scores over time (*β* = −6.93, p = 5.95×10^−06^). Life stress was a significant predictor of more severe EMA scores (*β* = 24.50, p = 9.99×10^−03^). MDD cases exhibited more inconsistent sleep patterns (*β* = 32.14, p = 5.44×10^−04^), shorter total sleep times (*β* = − 94.38, p = 2.82×10^−03^), and a higher frequency of naps (*β* = 14.05, p = 4.02×10^−03^). MDD cases took fewer steps per day (mean = 5828.64, sd = 6188.85) than controls (mean = 7088.47, sd = 5378.18) over the course of the study, but this difference was not significant (*p* = 0.33), activity levels were not significantly predictive of EMA score (*p* = 0.75).

**Conclusions:** This study demonstrates the feasibility of integrating smartphone-based EMA with wearable activity tracking in adolescents with depression. High compliance rates support the practicality of this approach, while EMA data provide valuable insights into the dynamic nature of depressive symptoms, particularly in relation to sleep and life stress. Future studies should validate these findings in larger, more diverse samples. Clinically, EMA and wearable tracking may enhance routine assessments and inform personalized interventions by capturing symptom variability and external influences in real time.

## Introduction

Adolescent depression is a significant mental health crisis, 14.7% of the adolescent population report at least one major depressive episode with severe impairment^1^. This trend predates the COVID-19 pandemic^2^ and has continued apace^3^. The impact of adolescent depression is severe, a depressive episode leads to immediate debilitating effects plus long-term consequences^4^, for example impaired academic performance^5^ and challenges in forming interpersonal relationships^6^. Depression that begins in adolescence often follows a recurrent pattern^7^, persisting into adulthood, and is associated with higher levels of anxiety, substance abuse, and impaired functioning in later life^8^. Importantly, depression does not manifest uniformly across individuals; Major Depressive Disorder (MDD) is known for its heterogeneous symptomatology^9^. The course and severity of depression can vary widely among individuals^10^. Moreover, depressive symptoms can fluctuate within the same individual over time^11,12^. Despite the fluctuating and dynamic nature of MDD, typical clinical assessment methods are cross-sectional. Most standard symptom assessments often require patients to recall how they have felt for the previous 1-2 weeks^13^. Recall of mood states in the previous weeks may be inaccurate depending on cognitive style and illness severity^14–18^. Ecological Momentary Assessment (EMA) bridges this gap. EMA refers to the repeated and brief assessments of the same person over multiple days. EMA enables a granular view of depressive symptoms in real-world settings, overcoming the temporal and contextual limitations of standard assessments^19^. In particular, EMA provides increased ecological validity by decreasing recall bias^20^. It also enables the measurement and modelling of within person variability since changes can be tracked over time. Thus, this tool facilitates the assessment of symptom fluctuations, offering insights into the dynamic nature of the disorder. In addition, repeated assessment of one or more variables (e.g., mood and sleep) allows an examination of covariation between variables over time^21^.

Thanks to growth in technology EMA can be conveniently completed using the ubiquitous smartphone, and can be integrated with wearables, such as activity trackers (e.g., FitBit) to monitor sleep and physical activity^19^.Despite the many advantages of EMA and its potential clinical utility^22–24^,literature on the topic is limited by sample type, and also EMA method and schedule intensity^25^. The present manuscript adds to the extant literature in a number of ways. First, most EMA mood studies have been conducted in adults^25,26^, while the present study focuses on adolescents with major depressive disorder. Second, many studies that focused on adolescents used phone-based EMA^27–31,31,32,32–34,34–36^ where interviewers called participants on a schedule and recorded responses, or paper-based EMA^37,38^ where participants fill in paper diaries on a schedule, leaving the utility of smartphone-based EMA, which we used in the present study, still mostly unknown^25,39^. To our knowledge only one study has measured mood over time in depressed patients using smartphone, app-based EMA, which comprised a single mood question administered over the course of six days^40^. Third, there is a notable gap in terms of moderate duration studies with high frequency checkins, the present study had two weeks of twice daily checkins. Previous studies have either been much shorter (e.g., four days^29^) or longer studies but with less frequent checkins (e.g., eight weeks of checkins with four day intervals^30,33^). **Table 1** shows details of cited studies. More data is needed to demonstrate that momentary assessment of depressive symptoms in adolescents using smartphone-based apps is possible. Additionally, that the collected data behaves as expected, that is depressed subjects show greater severity and variability of symptoms than healthy controls, and that related measures covary with those symptoms.

**Table 1.** Characteristics of existing studies of ecological momentary assessment of mood and associated variables in adolescent samples with and without major depressive disorder.

| Study | Sample | Age Range (years) | EMA Focus | EMA Duration | Check-in Frequency | EMA Method | Actigraphy |
| --- | --- | --- | --- | --- | --- | --- | --- |
| **Axelsson<sup>27</sup>** | 5 controls<br>11 MDD<br>5 BP | 10-17 | Location, social context, mood, media use, future plans, significant events | Five four-day blocks (Fri-Mon) | 12 calls per block | Answer-only cellular phone | No |
| **Silk<sup>32</sup>** | 16 controls<br>19 MDD | 8-17 | Positive and negative affect, and companions | Five four-day blocks (Fri-Mon) | 12 calls per block | Answer-only cellular phone | No |
| **Whalen<sup>36</sup>** | 23 controls<br>30 MDD | 7-17 | Positive and negative affect, caffeine consumption | Five four-day blocks (Fri-Mon) | 12 calls per block | Answer-only cellular phone | No |
| **Mor<sup>38</sup>** | 278 healthy individuals | Mean = 16.9 | Mood, stress, self-focus, social activity | Three days | 6 per day | Paper diary | No |
| **Forbes<sup>29</sup>** | 28 controls<br>15 MDD | 8-17 | Positive and negative affect | Four days | 1 per day | Cell phone | No |
| **Silk<sup>33</sup>** | 32 controls<br>47 MDD | 7-17 | Behavior, emotion, social context | Five four-day blocks (Fri-Mon) | 12 calls per block | Answer-only cellular phone | No |
| **Cousins<sup>28</sup>** | 23 controls<br>23 anxiety<br>42 MDD | 8-16 | Positive and negative affect | Two four day blocks | 12 calls per block | Answer-only cellular phone | Yes |
| **Primack<sup>31</sup>** | 60 controls<br>46 MDD | 7-17 | Media exposures (e.g., internet) | Five four-day blocks (Fri-Mon) | 12 calls per block | Answer-only cellular phone | No |
| **Forbes<sup>30</sup>** | 31 MDD and anxiety<br>23 anxiety<br>12 MDD | 8-16 | Positive and negative affect, and companions | Four days | 12 calls total | Answer-only cellular phone | No |
| **Waller<sup>34</sup>** | 31 controls<br>29 MDD | 11-17 | Behavior, emotion, social context | Three five-day blocks (Thurs-Mon) | 42 calls total | Answer-only cellular phone | No |
| **Frost<sup>37</sup>** | 353 healthy individuals | 11-18 | Feelings and activities | Seven days | 8 per day | Paper diary | No |
| **Bickham<sup>75</sup>** | 125 healthy individuals | 12-15 | Media use | Two weeks | 48 assessments | Handheld computer | No |
| **Cushing<sup>76</sup>** | 20 healthy individuals | 13-18 | Affect and energy | Twenty days | 4 assessments per day | Cell-phone app on study device | No |
| **Minch<sup>40</sup>** | 253 healthy individuals | 12-17 | Mood | Six days | 30 assessments total | Text message | No |

The present study had three primary goals, all of which contribute to assessing the feasibility of smartphone-based EMA in adolescents with depression. First, we aimed to evaluate whether participants could be retained in the study and engage consistently with smartphone-based EMA and wearable data collection. Second, we assessed whether mood reports captured expected symptom variation, specifically whether adolescents with depression demonstrated greater severity and variability of mood and anxiety symptoms compared to healthy controls, supporting the validity and potential clinical utility of this method. Third, we examined whether participants found the app-based EMA approach acceptable by evaluating self-reported satisfaction and usability of the EMA methods. By addressing these key aspects of feasibility, this study provides critical insights into the viability of smartphone-based EMA for real-time symptom monitoring and personalized intervention development in adolescent depression. As this is a pilot study with a small sample size, the findings should be viewed as preliminary. Future studies with larger, more diverse samples will be necessary to confirm and expand upon the relationships observed here, including potential subgroup-specific patterns within the heterogenous construct of MDD.

## Methods

### Procedures

**Figure 1** depicts the study procedures. Study procedures were identical across participants regardless of case status. Participants completed the protocol across a mean time span of 7 weeks.

**Figure 1.**
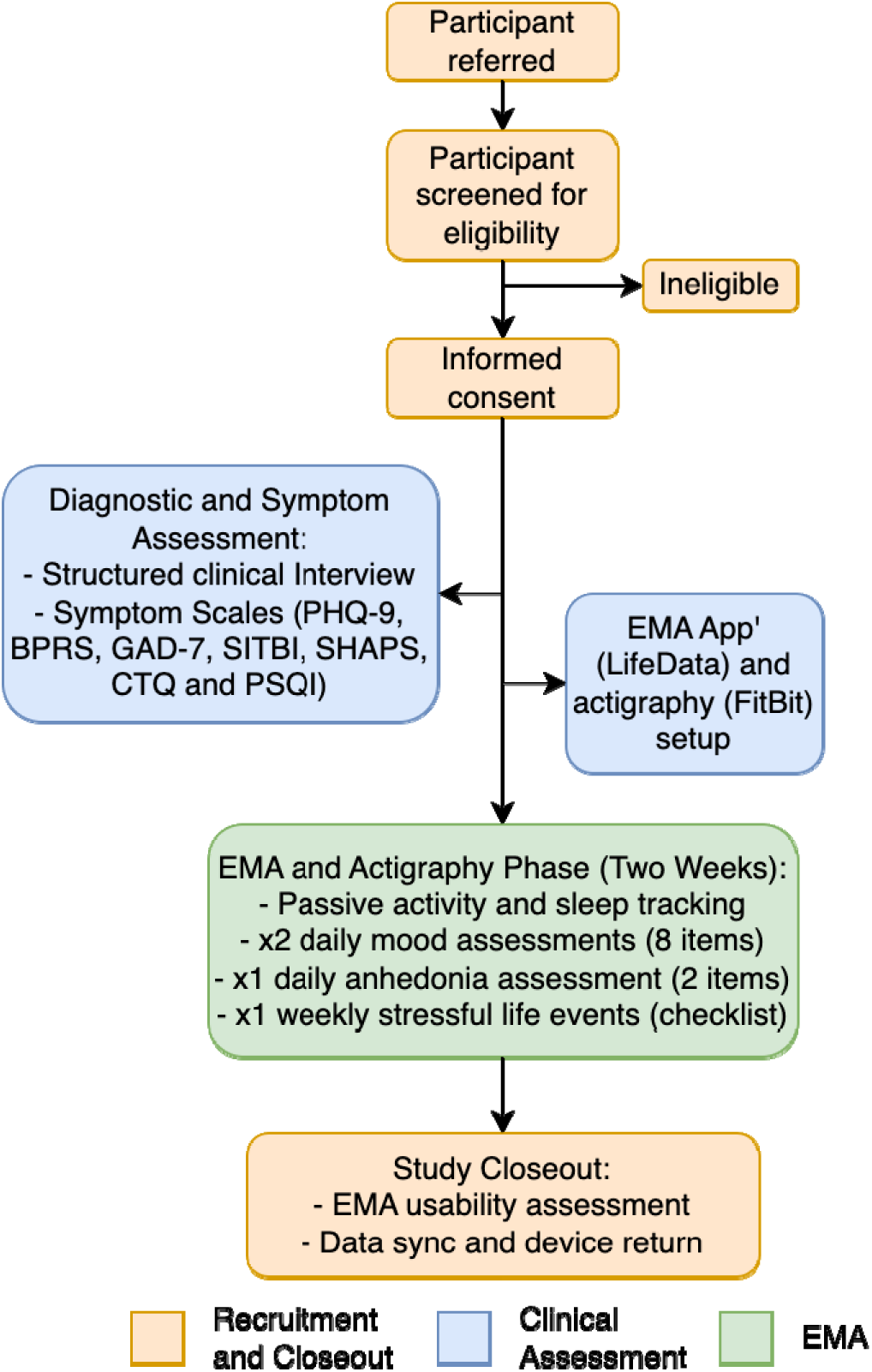
Study protocol. After being consented the presence (or absence) of major depressive diagnoses (and other DSM-V diagnoses) were confirmed using a structured clinical interview. Symptom scales for depression, general psychopathology, anxiety, suicidality and self-injury, anhedonia and trauma were administered. A smartphone app that enabled EMA to take place (LifeData) was setup on the participant’s phone. Each participant was given a FitBit and the device was linked with the participant’s smartphone (for data syncing). Participants completed two weeks of EMA and actigraphy concurrently. At the end of the two weeks participants returned the FitBit device and they completed a EMA usability assessment.

### Participant Recruitment

The sample comprised 36 individuals (75% female, mean age = 19.50 years, sd = 3.92 years, range = 14-27 years) recruited from the Boston, MA, area (**Table 2**). The majority of the sample identified as white (67%) with two individuals identifying as Hispanic or Latino, 4 as Black or African American and 1 as Asian. Twenty-three individuals had a major depressive disorder (MDD) and were recruited from Outpatient Psychiatry Services (OPS) at Boston Children’s Hospital. Providers requested for consent from the participant and their family for the research team to contact them about the study and research staff followed up to explain the study and, if appropriate, arrange for a consent visit to take place. The control group (N = 13) were recruited from the community via flyers and advertisements placed in Boston. Exclusion criteria for individuals with MDD included a severe neurodevelopmental disorder or other impairment that impacts the participants ability to provide the required information for the study (e.g., symptom reports), a substance or medication induced affective disorder, an affective disorder secondary to a medical condition, and a current Axis I psychotic or bipolar disorder. Exclusion criteria for controls included a severe neurodevelopmental disorder or a current or past psychiatric diagnosis as defined by DSM-V criteria.

**Table 2.**
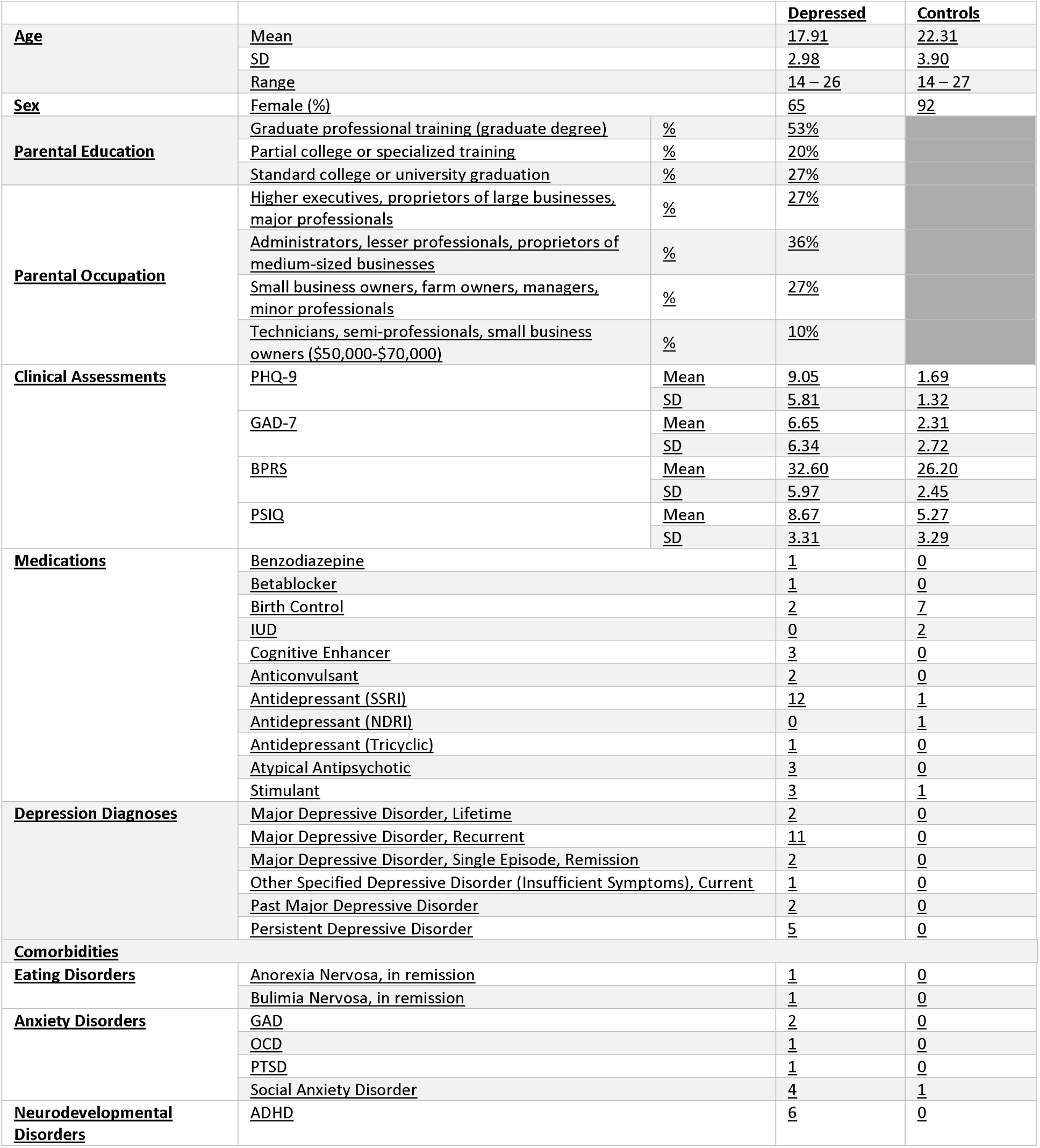
Baseline characteristics of the depressed and control groups.

### Ethical Considerations

This study was reviewed and approved by the Boston Children’s Hospital Institutional Review Board (BCH IRB). The study involved the collection and analysis of data from human subjects, and all procedures were conducted in accordance with the IRB-approved protocol. All participants provided informed consent/assent using forms approved by the institutional review board at Boston Children’s Hospital. This ensured that participants were fully informed about the study’s purpose, procedures, potential risks, and benefits before agreeing to participate. To maintain participant confidentiality, all collected data were stored in secure databases and de-identified prior to analysis. All data included in the present manuscript or any supplementary materials are completely anonymous. There was no cost or fee associated with study participation. Participants were compensated for their time and effort in completing study procedures. They were eligible to receive up to $144, with payments provided via ClinCard, a secure electronic payment system. Payment breakdown was as follows: $20 for SCID; $20 for questionnaire completion; $14 ($1 per day) for FitBit wearing; $14 for FitBit return; $14 for usability questionnaire; and $56 ($2 per check-in for EMA). Participants were paid for all aspects of the study that they completed, including individual EMA check-ins. No identifiable images of individual participants are included in this manuscript or supplementary materials.

### Diagnostic and Symptom Assessment

The presence or absence of major depression diagnoses (and comorbid disorders) were confirmed using the SCID^41^ (with additional modules from the KSAD interview^42^ if <18 years of age) in order to confirm past or current MDD (and other DSM diagnoses). Depressive symptoms were assessed at study intake using the Patient Health Questionnaire (PHQ-9^43^). Participants were also administered the Brief Psychiatric Rating Scale (BPRS^44^), Generalized Anxiety Disorder (GAD-7^45^), and the Pittsburgh Sleep Quality Index (PSQI^46^). Parents of probands were administered the Hollingshead Index of Socioeconomic Status^47^, 25 parents agreed to complete the scale.

### Ecological Momentary Assessment (EMA)

EMA data was recorded using LifeData^48^. LifeData is a smartphone-based EMA program that runs on both Android and iPhone smartphones. Check-ins are delivered via the LifeData mobile app RealLifeExp. EMA lasted two weeks, during this time participants received a push notification twice per day (6am and 4pm), reminding them to complete a check-in. Notifications were sent four times (at 1 hour intervals) per check-in allowing each participant a four-hour window to complete the check-in (6-10am and 4-10pm).

For each check-in participants were asked the same eight questions. Participants rated the items using a slider which translated to a score (0-100): “I am sad.”; “I feel bothered by every little thing.”; “I have no interest in things I would usually enjoy (e.g., food, TV, games, spending time with friends/family).”; “I do not have enough energy to get going.”; “I have no appetite or feel much hungrier than usual.”; “I feel physically tense and/or jittery.”; “I am nervous, anxious or on edge.”; and “I can’t stop worrying.” Items were designed to be reflective of established measures of depression (PHQ-9^43^) and anxiety (GAD-7^45^) with the adaptation that they could be administered multiple times per day (see **Table S1** for an explanation of each item). For a subset of statistical models (see below) the first five items were classed as measuring mood and the rest as measuring anxiety. Once per week, during an afternoon check-in, participants completed a checklist of stressful life events, the question reads “Did any of the following happen to you since the last time you completed one of these surveys? Check all that apply”. The participant selects as many of the following options as appropriate : “I argued with a friend or family member.”; “I was not allowed to do something I wanted to do.”; “I got a bad grade in school.”; “My parents have been arguing a lot.”; “Somebody in my family got a serious illness.”; “Someone in my family was arrested.”; “Somebody teased or threatened me.”; “I teased somebody else.”; “I did something that made me feel embarrassed.”; “Someone commented negatively on the way I look.”; “I was excluded from a group event.”; “I got disciplined or suspended from school.”

### Physical Activity (Steps) and Sleep Features

Participants were issued with a FitBit Charge 3 and were instructed to wear it for the duration of the EMA portion of the study. Twenty-two individuals (11 controls (mean age = 22.90, sd = 3.27 years, 10 females) and 11 MDD cases (mean age = 18.20, sd = 2.86 years, 7 females) wore the FitBit with sufficient regularity for data to be analyzed. Data were collected on daily activity (number of steps) and sleep.

Fitbit data were obtained via the Fitbit API, which provides preprocessed JSON files reflecting proprietary algorithms for step counting and sleep^49^. For sleep each participant had one JSON file that spanned the duration of the study which included a sleep log with timestamps for sleep onset and offset and a breakdown of sleep stages (e.g., light, REM, deep). For steps each JSON file contained minute-level step count data linked to a timestamp. Data in the JSON files were converted to csv using a combination of standard and pandas^50^ libraries in PyCharm^51^ (python version 3.9) and rearranged for analysis using tidyr^52^ and dplyr^53^ in R^54^. Following others work^55^ in the field we extracted numerous sleep features from the data collected using the Fitbit device. These related to sleep architecture, quality, and stability (see **Table S2** for details or individual metrics). A nap was defined as time spent in bed lasting < 180 minutes, FitBit does not calculate deep, light and REM sleep during these shorter sleeps. Therefore, naps were included in analyses focused on total sleep time (TST) and time in bed (TIB) but were excluded for analyses focused on sleep stage. Sleep quality metrics were calculated per all logged sleeps per individual, sleep stability metrics were derived across all logged sleeps. Steps that were recorded during a documented period of sleep were recoded to 0 as they were likely due to movement in bed rather than steps.

### Usability/Tolerability Questionnaire

At the end of participation all participants completed a short questionnaire that evaluated their experience using the EMA app (for example, if the app was easy to use; see Supplemental Material for a list of questions). All but one of the statements were rated from 1-7 with 1 being strongly disagree, 4 neutral and 7 strongly agree. Responses were recoded as disagree (1-3) and agree (5-7). Analysis included frequencies and proportions of responses per item. One item, relating to where participants used the app, included a free text response, unique responses are included in the supplemental materials.

### Statistical Analyses

All analyses were conducted in R (version 4.2.1)^54^.

### How does depressive symptomatology vary over time?

In line with others EMA work^56^ we opted to examine variability in depressive symptoms both visually and statistically. Time series data were examined using standardized (group-mean centered) and raw data. Group-mean centered plots depict the relative variability over time across the sample. Before plotting data were group-mean centered (by subtracting each participant’s mean from each individual score), which results in standardization of the figure such that units are in units of deviation from each individual’s average (set to 0). Per subject raw score time-series plots depict the fine-grained differences between each individual’s data. Data were plotted using ggplot2^57^.

We used several statistics to quantify variability including the root mean square of successive differences (RMSSD)^58^, coefficient of variation (CoV)^59^ and intraclass correlations (ICC). The RMSSD reflects variability over time. The CoV reflects the dispersion of the data and is useful for comparing variability when means differ (e.g., in cases versus controls). The ICC reflects the proportion of variance attributable to between-person variability, thus 1-ICC indicates the proportion of variability attributable to within-person variability. Variability statistics were calculated using a combination of base R and the psych^60^ package.

### Predictors of Depression Severity and Variability Over Time

The EMA data were non-independent, for example we might expect data collected within the same person to be more strongly related than those collected from two different people. This structure is sometimes referred to as multilevel, observations are nested within item type (mood or anxiety), which are nested within time of day (AM or PM) and day, which are nested within participant. Therefore, when evaluating the impact of various predictors on EMA scores we applied linear mixed-effects regression to the data using the package lme4^61^. Effect sizes for fixed effects (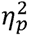) were calculated using the effectsize package^62^, and were interpreted as small effect: 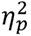 ≈ 0.01, medium effect: 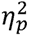 ≈ 0.06, large effect: 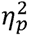 ≈ 0.14. Similarly, observations for sleep architecture and quality were nested within day (where some individuals logged sleeps with onset times within the same day), within sleep stage, within participant. Multilevel modeling is robust to missing data and therefore we did not impute the small amount of missing data that was present in the EMA, of 1008 expected check-ins 923 were observed (overall compliance = 92%). Prior to testing the effect of various predictors on depression we tested an intercept only, or unconditioned, model to evaluate the need for random effect). Significant random effects (determined using the lmerTest^63^ package) were retained in subsequent models. Given the different distributions of age and sex in case and control groups, these variables were included as covariates in all models.

First, we asked whether individuals with a MDD diagnosis demonstrated greater symptom severity (indexed using EMA score) and variability than controls. We also evaluated the impact of sex and age on severity as well as fixed effects of Time of Day and Item Type. Second, we examined whether anxiety and mood covaried over time and whether anxiety or mood the preceding check-in significantly predicted mood the following one. Third, we asked whether life stress was associated with increased symptom severity. Fourth, we examined whether activity levels (number of steps) were associated with MDD diagnosis or symptom severity. Fifth, we assessed whether number of minutes asleep, sleep onset, or sleep waking was associated with MDD diagnosis or symptom severity. Sixth, we examined the results of the usability questionnaire.

## Results

The total unique check-ins completed by participants = 923 (mean check-ins per participant = 25.60, sd = 2.78). Assessments were completed across a total of 486 unique days (mean days per participant = 13.50, sd = 1.25), and yielded 1.90 mean check-ins per participant per day. For the two required check-ins per day compliance rate = 91.57%, cases demonstrated slightly lower compliance (89.94%) than controls. Across individual EMA items this yielded 7336 data points for regression modeling.

### How does depressive symptomatology vary over time?

**Table 3** shows the descriptive and variability statistics for the Sum Score of all EMA items as well as for the individual items. In terms of reported symptom severity, MDD cases scored higher than controls on all items. Examination of variability statistics (**Table 3**) indicate that there was considerable variability in both the sum score and the individual items. The RMSSD and CoV suggest greater variability in MDD cases than in controls. Examination of the ICC across items suggests that ∼50% of the variance in item-level responses was due to within-person variance in MDD cases.

**Table 3.**
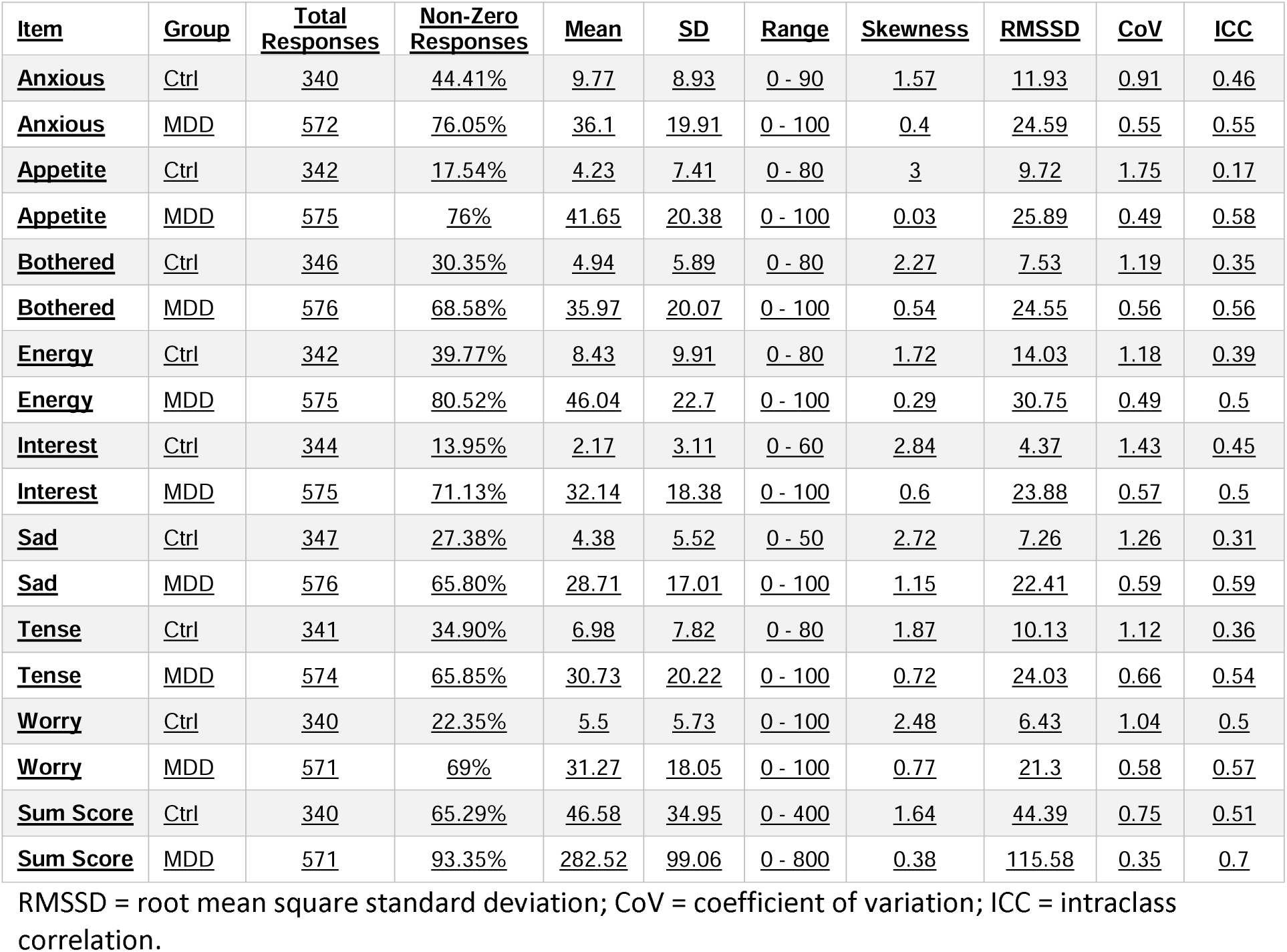
Descriptive and Variability Statistics for individual ecological momentary assessment items in depressed and control groups.

| **Item** | **Group** | **Total Responses** | **Non-Zero Responses** | **Mean** | **SD** | **Range** | **Skewness** | **RMSSD** | **CoV** | **ICC** |
| --- | --- | --- | --- | --- | --- | --- | --- | --- | --- | --- |
| **Anxious** | Ctrl | 340 | 44.41% | 9.77 | 8.93 | 0 - 90 | 1.57 | 11.93 | 0.91 | 0.46 |
| **Anxious** | MDD | 572 | 76.05% | 36.1 | 19.91 | 0 - 100 | 0.4 | 24.59 | 0.55 | 0.55 |
| **Appetite** | Ctrl | 342 | 17.54% | 4.23 | 7.41 | 0 - 80 | 3 | 9.72 | 1.75 | 0.17 |
| **Appetite** | MDD | 575 | 76% | 41.65 | 20.38 | 0 - 100 | 0.03 | 25.89 | 0.49 | 0.58 |
| **Bothered** | Ctrl | 346 | 30.35% | 4.94 | 5.89 | 0 - 80 | 2.27 | 7.53 | 1.19 | 0.35 |
| **Bothered** | MDD | 576 | 68.58% | 35.97 | 20.07 | 0 - 100 | 0.54 | 24.55 | 0.56 | 0.56 |
| **Energy** | Ctrl | 342 | 39.77% | 8.43 | 9.91 | 0 - 80 | 1.72 | 14.03 | 1.18 | 0.39 |
| **Energy** | MDD | 575 | 80.52% | 46.04 | 22.7 | 0 - 100 | 0.29 | 30.75 | 0.49 | 0.5 |
| **Interest** | Ctrl | 344 | 13.95% | 2.17 | 3.11 | 0 - 60 | 2.84 | 4.37 | 1.43 | 0.45 |
| **Interest** | MDD | 575 | 71.13% | 32.14 | 18.38 | 0 - 100 | 0.6 | 23.88 | 0.57 | 0.5 |
| **Sad** | Ctrl | 347 | 27.38% | 4.38 | 5.52 | 0 - 50 | 2.72 | 7.26 | 1.26 | 0.31 |
| **Sad** | MDD | 576 | 65.80% | 28.71 | 17.01 | 0 - 100 | 1.15 | 22.41 | 0.59 | 0.59 |
| **Tense** | Ctrl | 341 | 34.90% | 6.98 | 7.82 | 0 - 80 | 1.87 | 10.13 | 1.12 | 0.36 |
| **Tense** | MDD | 574 | 65.85% | 30.73 | 20.22 | 0 - 100 | 0.72 | 24.03 | 0.66 | 0.54 |
| **Worry** | Ctrl | 340 | 22.35% | 5.5 | 5.73 | 0 - 100 | 2.48 | 6.43 | 1.04 | 0.5 |
| **Worry** | MDD | 571 | 69% | 31.27 | 18.05 | 0 - 100 | 0.77 | 21.3 | 0.58 | 0.57 |
| **Sum Score** | Ctrl | 340 | 65.29% | 46.58 | 34.95 | 0 - 400 | 1.64 | 44.39 | 0.75 | 0.51 |
| **Sum Score** | MDD | 571 | 93.35% | 282.52 | 99.06 | 0 - 800 | 0.38 | 115.58 | 0.35 | 0.7 |

RMSSD = root mean square standard deviation; CoV = coefficient of variation; ICC = intraclass correlation.

**Figure 2** shows the group-mean centered time series plot of the Sum Score. At the group level, scores fluctuated in a saw-tooth pattern, with repeated rises and falls but no clear linear trend of improvement or worsening over time. This pattern reflects short-term variability in EMA data and highlights overall fluctuations in symptom severity across the sample. Item-level data followed a similar pattern (**Figure S1**), in MDD cases variability in symptom reporting did not appear to vary as a function of item. Controls appear to report a greater degree of variability to the irritability and anhedonic items (Bothered and Interest) than they did on mood or anxiety symptoms.

**Figure 2.**
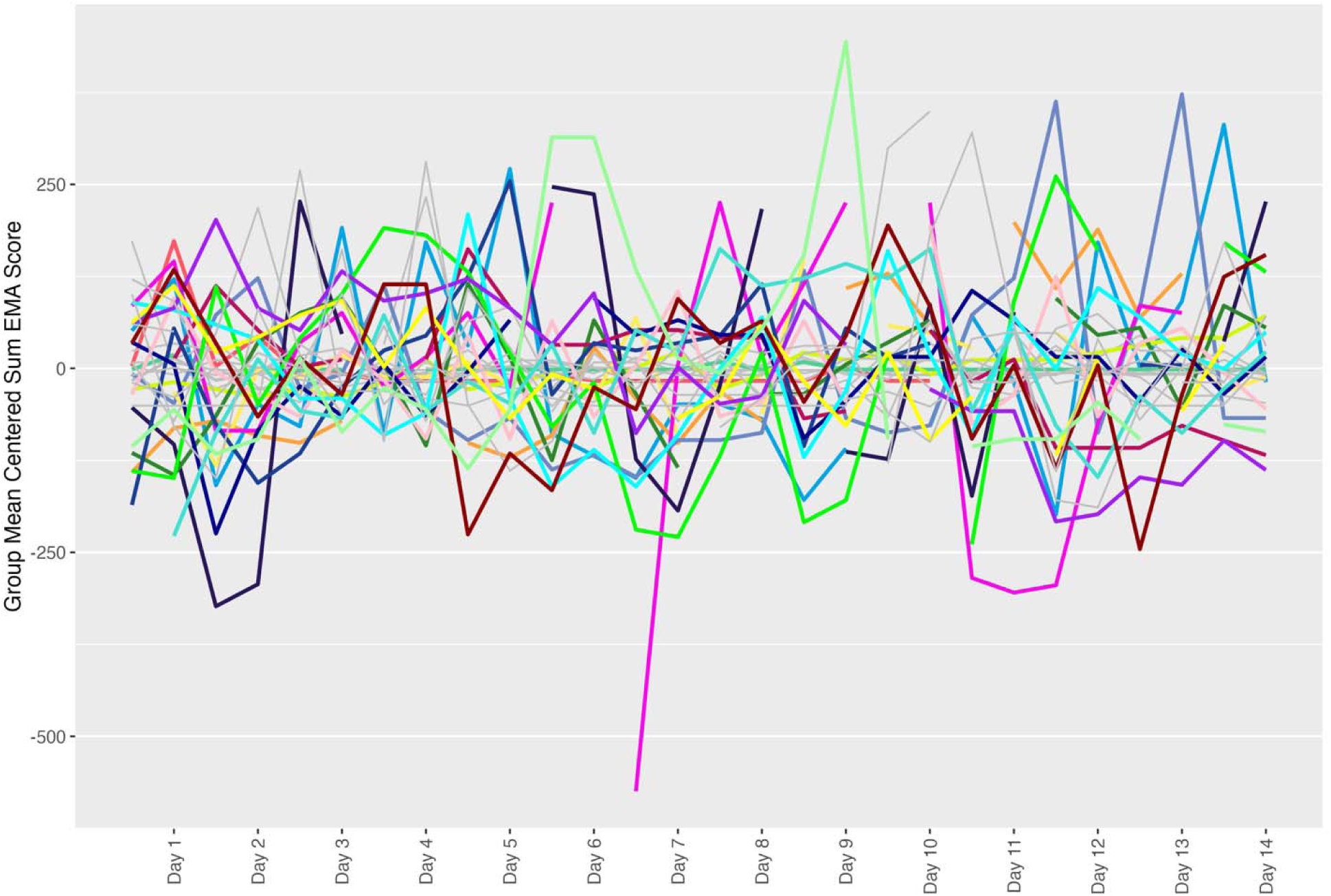
Time-series plot of the group-mean centered EMA data (sum score across all eight items). Colored lines represent individuals with a MDD diagnosis, grey lines represent controls.

At the individual level we observe heterogeneity (Figure 3) where some individuals with a MDD diagnosis demonstrate worsening throughout the course of the study (e.g., 30001, 30015, 30028, 30030, 30044, 30048), and others appear to improve (e.g., 30035, 30036, 30047). Interestingly, not all controls demonstrated floor effects. While controls reported more zero responses than cases (**Table 3**) some endorsed minimal symptoms (e.g., 30011, 30033, 30038, 30046, 30057) throughout the course of the study; admittedly these symptoms would likely amount to sub-threshold symptoms for a MDD diagnosis but this suggests that healthy controls can demonstrate variation in depressive symptomatology. Three MDD cases (30003, 30012 and 30019) demonstrated minimal symptoms; diagnoses for these individuals were in line with low symptoms (Other Specified Depressive Disorder (Insufficient Symptoms), Recurrent Major Depressive Disorder and Past Major Depressive Disorder Single Episode respectively).

**Figure 3.**
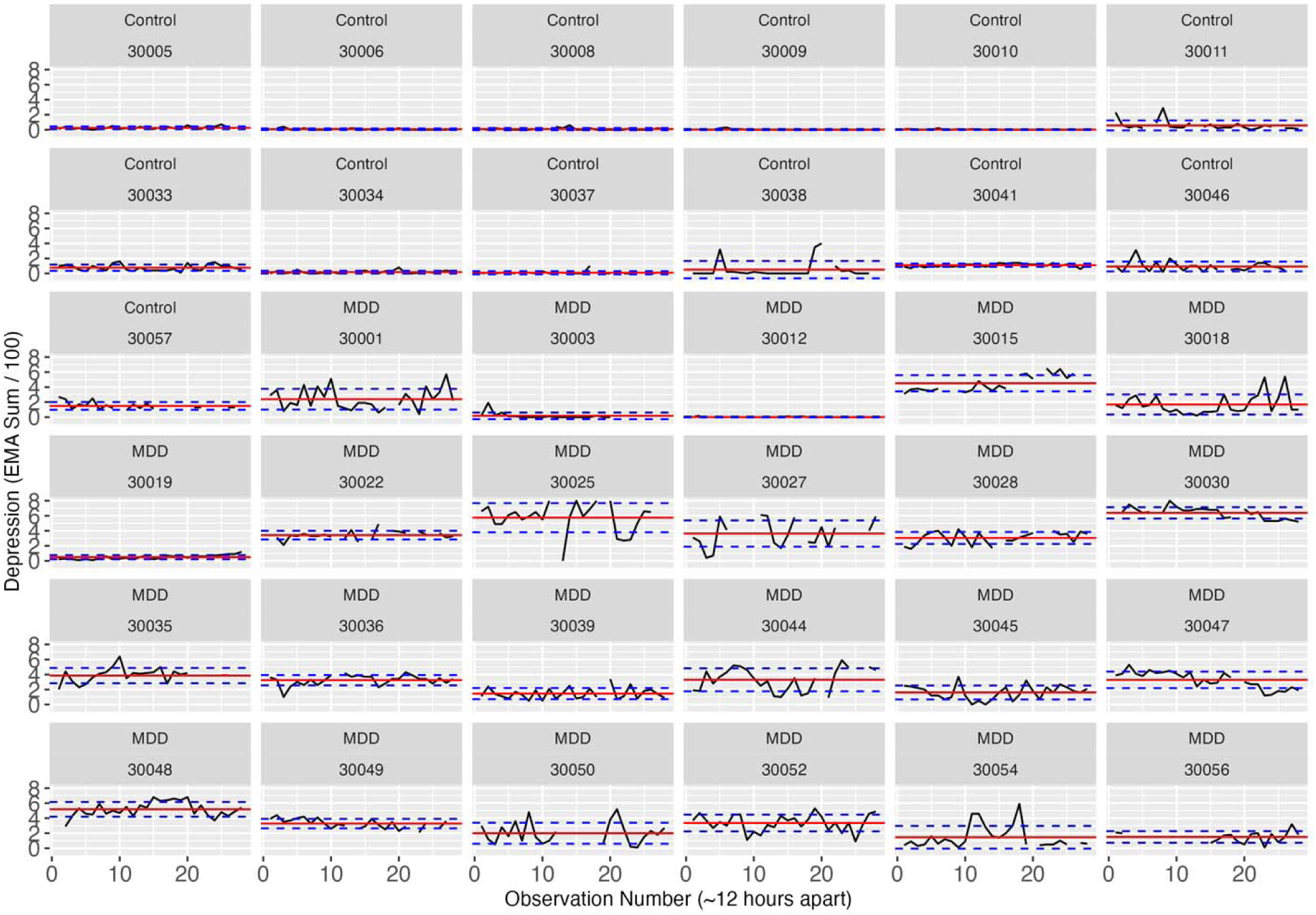
Time series plots of the raw EMA data (sum score across all eight items). The black line represents the reported score, dashed lines are controls and solid lines are those with a MDD diagnosis. Mean score (per individual) is represented by the red horizontal line and blue dashed lines represent +/I 1 SD.

### Predictors of Depression Severity and Variability Over Time

The unconditioned model indicated significant between-subject nested effects of Participant, Day and Item Type but not Time of Day (**Table S3**). All significant random effects were retained in subsequent modeling.

#### 1. Effect of MDD on EMA Score Severity Over Time

We examined the impact of MDD diagnosis on summed EMA score using a linear mixed model (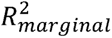 = 0.26, 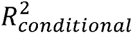 = 0.65) with fixed effects of MDD (plus age, sex, item type, and time of day). The model indicated significant main effects of MDD (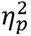 = 0.41, *β* = 34.48, 95%CI[20.22, 48.74], *p* = 2.17×10^−06^), sex (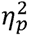 = 0.16, *β* = 16.81, 95%CI[3.55, 30.07], *p* = 1.29×10^−02^), and a small but significant effects of item type (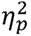 = 0.01, *β* = −1.69, 95%CI[−2.27, −0.55], *p* = 1.36×10^−02^) and time of day (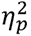 = 0.002, *β* = −1.41, 95%CI[−2.27, −0.55], *p* = 1.36×10^−03^). The effect of age (*p* = 0.85)was not significant.

We fit an additional linear mixed model (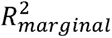 = 0.27, 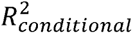 = 0.65) with MDD*time of day and MDD*item type interactions to establish whether there was greater diurnal variation in mood in cases than controls and whether there was a greater effect of item type in cases than in controls. There was a significant MDD*time of day interaction (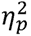 = 0.001, *β* = −2.54, 95%CI[−4.32, −0.76], *p* = 5.14×10^−03^; **Figure S2**) where cases exhibited diurnal mood variation (higher scores in the morning) but controls did not, 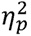 indicates that the effect was minimal. There was also a significant MDD*item type interaction (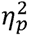 = 0.001, *β* = −6.93, 95%CI[−9.64, −4.21], *p* = 5.95×10^−06^; **Figure S3**) where cases demonstrated worse mood than anxiety scores and controls demonstrated the opposite effect, however 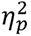 indicates that the effect was minimal.

PHQ-9 score at baseline (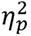 = 0.58, *β* = 2.74, 95%CI[1.92, 3.57], *p* = 8.82×10^−11^) significantly predicted EMA scores over the course of the study, suggesting that the items we selected for EMA have good convergent validity with an established measure of depression.

#### 2a. Cross-Sectional Covariation of Mood and Anxiety Over Time

We examined cross-sectional relationships between mood and anxiety over time using a linear mixed model (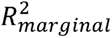 = 0.33, 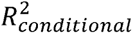 = 0.87) of summed EMA mood items with fixed effects of summed EMA anxiety items and MDD (plus age and sex). This model indicated significant covariation between mood and anxiety over time (**Figure S4** shows raw data per participant), main effects of anxiety (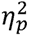 = 0.25, *β* = 0.64, 95%CI[0.56, 0.71], *p* = 2.70×10^−59^) and MDD (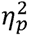 = 0.45, *β* = 132.14, 95%CI[81.24, 183.05], *p* = 4.26×10^−07^) were significant, but neither age (*p* = 0.22) nor sex were (*p* = 0.39).

#### 2b. Impact of Preceding Anxiety Score on Mood

We examined whether prior anxiety ratings predicted later mood scores using a linear mixed model with the same predictors as above plus anxiety score from the preceding check-in (where T = time, T-1) as a fixed effect (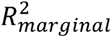 = 0.63, 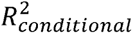 = 0.89). The preceding anxiety score was a nominally significant fixed effect but the size of the effect was small (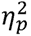 = 0.01, *β* = 0.11, 95%CI[0.04, 0.19], *p* = 09×10^−03^).

#### 2c. Impact of Preceding Mood on Subsequent Mood Score

We examined whether prior mood score predicted later mood score using a linear mixed model (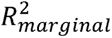 = 0.46, 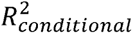 = 0.88) with T-1, T-2 and T-3 mood scores as a fixed effect in addition to MDD (plus age and sex). The immediately preceding mood score (T-1; 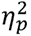 = 0.02, *β* = 0.11, 95%CI[0.04, 0.18], *p* = 2.73×10^−03^) was a significant predictor of subsequent mood score, as was mood score that was separated by one check-in (T-2; 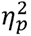 = 0.03, *β* = 0.16, 95%CI[0.09, 0.23], *p* = 2.72×10^−05^) and two (T-3; 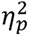 = 0.02, *β* = 0.11, 95%CI[0.04, 0.18], *p* = 1.74×10^−03^) check-ins. MDD (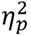 = 0.53) remained a significant predictor of mood score, age (*p* = 0.66) and sex (*p* = 0.55) were not significant predictors in this model.

#### 3. Effect of Life Stress on EMA Score Severity

Both MDD cases and controls reported life stress (**Table S4**). Most stressful life events in controls related to arguments with friends or family and embarrassment. These stressors were also common in MDD cases but they also endorsed stress related to instances of not being allowed to do what they wanted, being teased (and teasing others) or having negative comments made about their appearance. MDD cases reported 91 stressful life events while controls reported 35. Because controls were older than MDD cases and because the type of events included in the checklist were focused on younger individuals differences in frequency of life stress is likely not meaningful. A linear model comparing the number of stressful life events between groups while covarying for age and sex indicated a non-significant effect of group (*p* = 0.43). Notwithstanding this confound, looking at the entire sample EMA scores were higher in individuals that reported life stress, this was true in both controls and MDD cases (**Figure S5**). We examined cross-sectional and lagged relationships between self-reported stress and summed EMA score across each week (stress was reported once per week) using linear mixed models, including MDD, age and sex as predictors. Concurrent self-reported stress, occurring in the same week, significantly predicted EMA scores (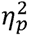 = 0.16, *β* = 24.50, 95%CI[6.07, 42.93], *p* = 9.99×10^−03^), whereas lagged stress (T-1; 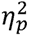 = 0.07, *β* = 6.15, 95%CI[−1.52, 13.82], *p* = 0.28) did not, suggesting an immediate rather than cumulative impact of stress () on mood and anxiety. MDD (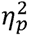 = 0.16, *p* = 5.05×10^−06^) and sex (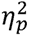 = 0.14, *p* = 2.82×10^−02^) remained significant predictors, age (*p* = 0.48) was not a significant predictor.

#### 4. Relationship Between MDD and Mood and Activity Levels

Number of steps taken per day (and across the course of the study) ranged widely between participants (**Figure S6**). MDD cases took fewer steps per day (mean = 5828.64, sd = 6188.85) than controls (mean = 7088.47, sd = 5378.18) over the course of the study. However, MDD was not a significant predictor of steps taken (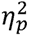 = 0.18, *p* = 0.33). We also examined cross-sectional relationships between EMA summed score and steps taken each day over time using a linear mixed model, steps were not a significant predictor of EMA score (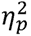 = −0.01, *p* = 0.75). Thus, activity, indexed using number of steps taken per day, and mood were not significantly related in the present study.

#### 5. Relationship Between MDD and Mood and Sleep

In both cases and controls, sleep onset was most commonly between 8pm and 4am (**Figure S7**) and sleep offset was most commonly between 5am and 11am (**Figure S8**), though there was more spread in sleep offset hour in cases. Descriptive statistics of sleep metrics are shown in **Table 4**.

**Table 4.** Descriptive statistics of sleep metrics in depressed and control groups.

|  |  | **Control** | **MDD** |
| --- | --- | --- | --- |
| **Nights** | **N** | **160** | **232** |
| **Individuals** | **N** | **11** | **11** |
| **Total Sleep Time (TST)** | Mean<br>(SD) | 388.62<br>(108.56) | 277.19<br>(163.31) |
| **Time in Bed (TIB)** | Mean<br>(SD) | 439.84<br>(124.20) | 315.11<br>(186.48) |
| **Time Awake in Bed (TAB)** | Mean<br>(SD) | 51.12<br>(20.95) | 37.41<br>(27.57) |
| **Deep Sleep** | Mean<br>(SD) | 79.24<br>(23.69) | 66.47<br>(27.90) |
|  | % | 17 | 15 |
| **Light Sleep** | Mean<br>(SD) | 264.05<br>(55.20) | 250.49<br>(82.77) |
|  | % | 57 | 58 |
| **REM Sleep** | Mean<br>(SD) | 94.80<br>(34.44) | 83.43<br>(39.12) |
|  | % | 20 | 19 |
| **Efficiency (TST/TIB)** |  | 0.89 | 0.89 |
| **5-Minute Wakes** |  | 390 | 370 |
| **Hypersomnia** |  | 2 | 5 |
| **Number of Naps** |  | 13 | 88 |
| **Coefficient of Variation (CoV) of TST** |  | 27.93 | 58.92 |
| **Mode Onset Hour** |  | 23:00 | 23:00 |
| **SD Onset Time (Minutes from 12pm)** |  | 123.54 | 202.99 |
| **Mode Offset Hour** |  | 7:00 | 06:00 |
| **SD Offset Time (Minutes from 12pm)** |  | 113.48 | 166.14 |

In terms of sleep staging, inspection of **Table 4** shows that the amount of deep, light and REM sleep between cases and controls did not differ, neither did sleep efficiency (how much time in bed was spent asleep). However, there were differences in (a) total sleep time (TST), (b) number of naps, and (c) variability in sleep length. We tested differences between cases and controls using linear mixed models, age and sex were included as covariates in all models. Cases spent significantly less time asleep (TST; 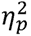 = 0.20, *β* = −94.38, 95%CI[−178.62, −10.14], *p* = 2.82×10^−03^) and time in bed (TIB; 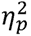 = 0.18, *β* = −103.39, 95%CI[−199.85, −6.94], *p* = 3.57×10^−02^). Cases took significantly more naps (*β* = 14.05, 95%CI[2.32 – 85.02], *p* = 4.02×10^−03^). Cases had significantly more variability in their total sleep time (the length of each sleep was more inconsistent than that taken by controls) as indexed by the CoV of TST (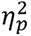 = 0.53, *β* = 32.14, 95%CI[16.04, 48.23], *p* = 5.44×10^−04^). We examined the relationship between TST and EMA data using a linear mixed model. The effect of TST on EMA score the following day was not significant (*p* = 0.69).

#### 6. Did participants find using the app to report on their mood acceptable?

**Figure 4** shows the results of the usability questionnaire. Inspection of Figure 4 shows that the majority agreed that the EMA app was easy to use (96%) and that they were comfortable using the app in social settings (77%); participants reported using the app in a wide variety of places (see **Table S5** for a complete list of unique responses). 100% of individuals found the app an acceptable way to report on their mood, 89% of them said they felt the app improved their access to research participation and 63% agreed that they would like to use an app like this to communicate with their healthcare provider.

**Figure 4.**
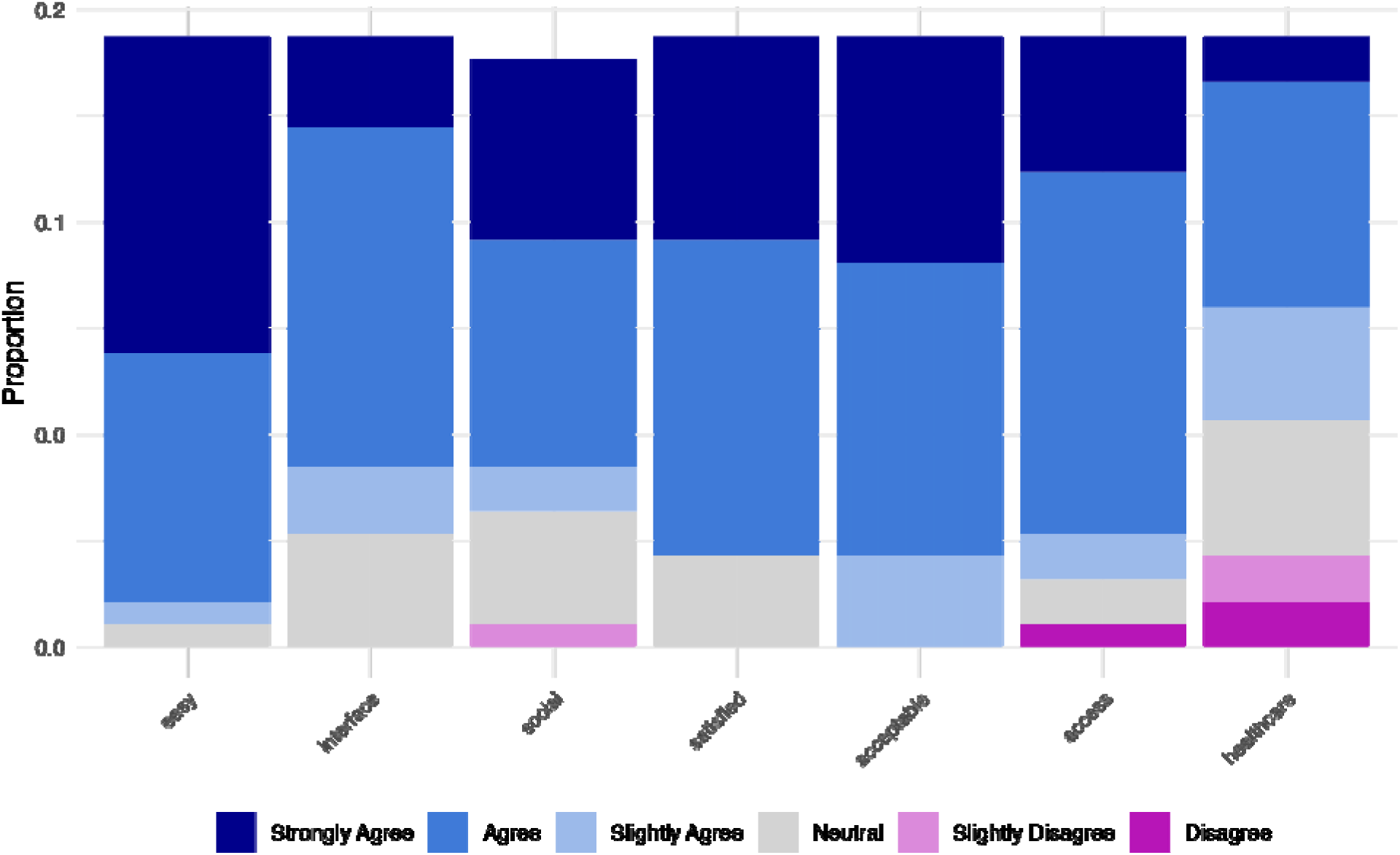
Usability data regarding the EMA app. The majority of participants agreed that the app was”easy” to use, that they liked the “interface” of the app, that they felt comfortable using the app in “social” settings, that they were “satisfied” with the app, and that the app provided them with an “acceptable” way to report on their mood. A small minority of participants disagreed that the app increased their “access” to research participation (N = 1) and that they would like to use a similar app to communicate with their “healthcare” provider (N = 4), but the majority were in agreement.

## Discussion

The present study comprehensively assessed the feasibility of smartphone-based EMA in adolescents with depression by (1) evaluating participant retention and engagement, (2) examining whether mood reports captured expected symptom variation, and (3) determining the acceptability of the app-based approach. Together, these findings demonstrate that smartphone-based EMA is a feasible, viable and effective method for real-time symptom monitoring in adolescents with depression.

Engagement with EMA was high, with exceptional compliance rates across participants. This suggests that adolescents were able and willing to complete frequent mood assessments over an extended period, reinforcing the feasibility of implementing this method in both research and clinical settings. The usability data indicate that participants found the app easy and satisfying to use, that they used the app wherever they were including in social settings, and that most participants would be amenable to using such an app in a clinical setting. These responses indicate that adolescents find smartphone-based apps for mood reporting convenient and further suggest that approaches using them, whether that be in the context of research or in the clinic, are likely to be well tolerated.

Beyond high compliance and self-reported acceptability, feasibility was further supported by the clinical distinctions captured in the collected data. Adolescents with depression reported significantly more severe and variable symptoms than healthy controls, reinforcing EMA’s ability to differentiate between clinical and non-clinical populations. This supports the discriminant validity of our approach, further reinforcing its utility and feasibility as a real-time assessment tool. The EMA scores were significantly predicted by baseline PHQ-9 scores, supporting its criterion validity against an established depression measure. While this alignment was expected given that EMA items were adapted from the PHQ-9 and GAD-7, it is notable that these adaptations retained the ability of those items to reflect depression severity while also being suitable for repeated, momentary assessment. Furthermore, individual-level trajectories revealed that while some participants experienced worsening symptoms, others improved. These findings underscore EMA’s ability to capture dynamic mood changes^9,22–24,64^ that would be difficult to assess with retrospective measures^14–19^. Notably, some healthy controls also reported transient depressive symptoms, underscoring the importance of including control groups in EMA studies and highlighting the potential for identifying individuals with emerging depressive episodes. Interestingly, increased life stress was associated with greater mood and anxiety symptom severity across both groups. The effect of stress on EMA scores was observed concurrently, but not lagged, indicating an immediate rather than cumulative effect of stress on mood and anxiety symptoms.

Activity data revealed that MDD cases spent less time asleep and in bed but took more naps than controls, suggesting that their sleep was more fragmented and variable than in controls. These findings are consistent with previous research showing an association between sleep disturbances and depression and anxiety^65,66^ particularly short sleep duration and symptoms of depressed mood^67^ and increased napping in adults^68,69^ and adolescents^70^ with depression. Although we did not find a significant relationship between daily mood variations and the previous night’s sleep or daily physical activity levels, our study supports the notion that sleep disturbances are a critical component of adolescent depression^71^.

The study should also be viewed in the context of some important limitations. First, our sample size limits the generalizability of our results. The sample size is small and so future replication of our main findings is key. Second, the racial homogeneity and relatively high SES of our sample, with 67% of participants identifying as White, may limit the generalizability of our findings to more diverse populations. Third, there is an age disparity between the studied groups, with the controls being significantly older on average than the MDD cases. This could have introduced an age-related bias, particularly for the age-specific assessments like the life stress questionnaire. Fourth, while fitbit is a cost effective way to collect sleep and activity information it may overestimate sleep time (particularly sleep staging estimates^72,73^) and activity levels^74^ compared to gold standard methods (e.g., polysomnography for sleep). We did not find differences between cases and controls in terms of number of steps taken or in terms of sleep staging, these might be true findings or the lack of difference might be clouded by the measurement error associated with Fitbit devices. Future research should aim to include a larger and more age-matched sample using alternate devices to mitigate these limitations.

When considering the clinical applications of EMA, it is important to acknowledge that this study was conducted as a paid research project. As a result, the high compliance rates observed may have been influenced by financial compensation, which does not necessarily translate to similar real-world clinical settings. Additionally, the implementation of EMA in clinical practice presents several challenges that must be addressed. These include logistical barriers, such as ensuring access to the necessary technology, as well as concerns about privacy and data confidentiality, particularly when working with vulnerable populations like children.

The findings of this study have strong implications for both research and, potentially, clinical practice. From a research perspective, this study is one of the first to explore the integration of smartphone-based EMA and wearable activity tracking in adolescents with depression. The findings highlight the potential of these technologies to capture real-time data and provide valuable insights into the dynamic nature of depressive symptoms. Future studies should explore the use of EMA and activity tracking methods in a larger and more diverse population to validate our findings and ultimately expand this novel method to clinical practices. In terms of clinical practice, the demonstrated feasibility of EMA and activity tracking in adolescents suggests that these methods could be integrated into routine clinical assessments to provide a more comprehensive and dynamic picture of symptomatology. Moreover, the development of personalized intervention strategies that take into account individual patterns of symptom fluctuations could lead to more effective management of adolescent depression^56^. A more comprehensive understanding of the external factors influencing symptom variability would provide insight into this relationship, particularly by looking at external factors such as environmental stressors and social interactions.

## Acknowledgements

This work is supported by a Tommy Fuss Independent Investigator Award (Grant Number: 92188)

## Data Availability

The data supporting the findings of this study are available upon request. Interested researchers may contact the corresponding author at for access.

