## supplemental_materials for "Pilot Study of Smartphone Ecological Momentary Assessment and Wearable Activity Tracking in Pediatric Depression"

**Methods**

***Usability/Tolerance Questionnaire***

Participants were asked to rate a series of statements regarding their experience using the LifeData app when completing the EMA portion of the study. These included: (1) The LifeData app was easy to use; (2) I like the interface of the app; (3a) I feel comfortable using this app in social settings; (3b) In which social settings did you use the app? (e.g., school, mall, at a friends house); (4) Overall, I am satisfied with this app; (5) This app provides an acceptable way for me to report on my mood; (6) The app improved my access to research participation; and (7) If it were possible, I would like to use an app like this to communicate with my healthcare provider. Each statement except for 3b was rated using radio button with the following options: Strongly Disagree; Disagree; Slightly Disagree; Neutral, Slightly Agree; Agree; and Strongly Agree. For item 3b a free text box was provided for participants to enter their responses.

**Results**

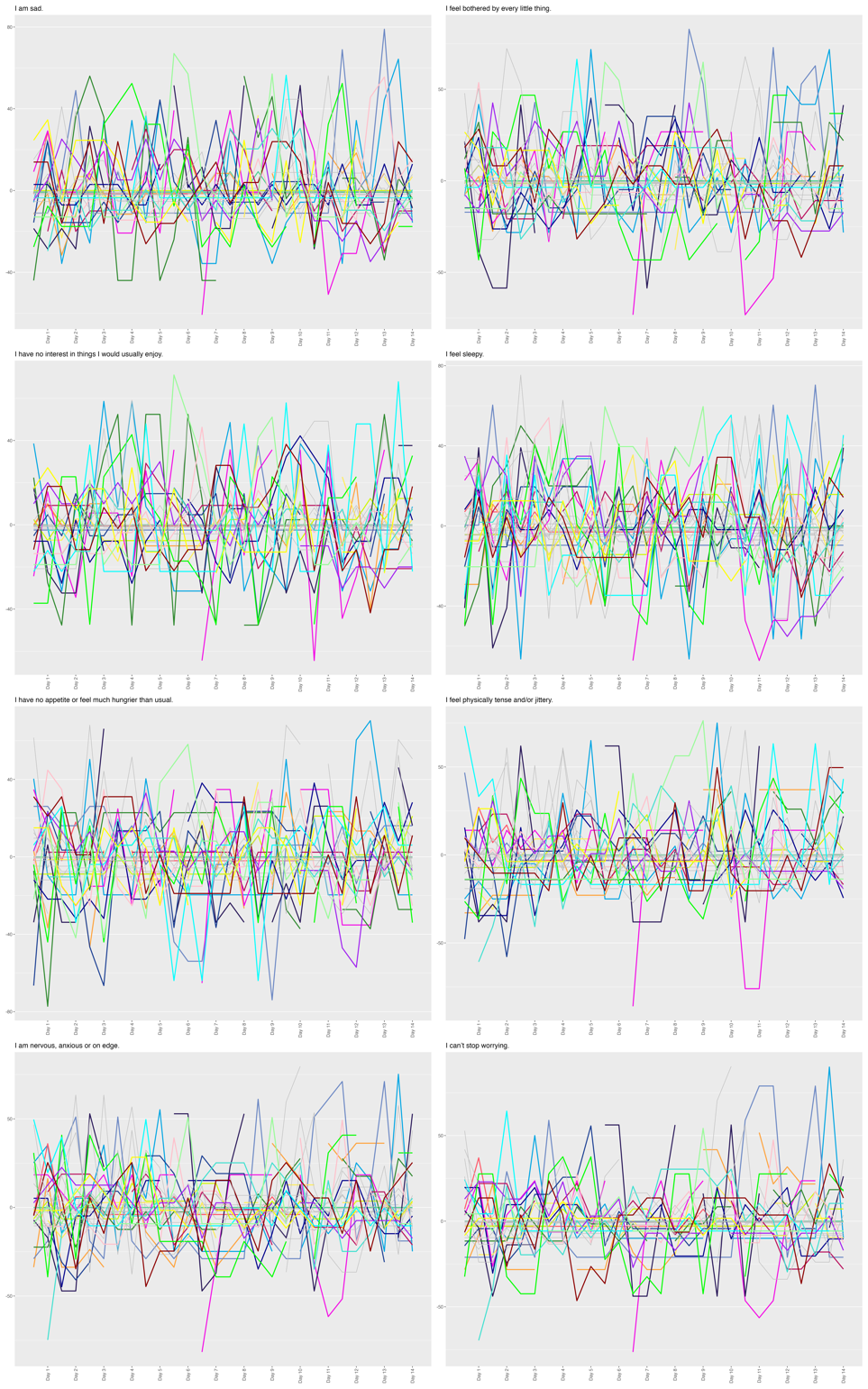

**Figure S1.** Time-series plots of group-mean centered individual EMA items. Colored lines represent individuals with a MDD diagnosis, grey lines represent controls.

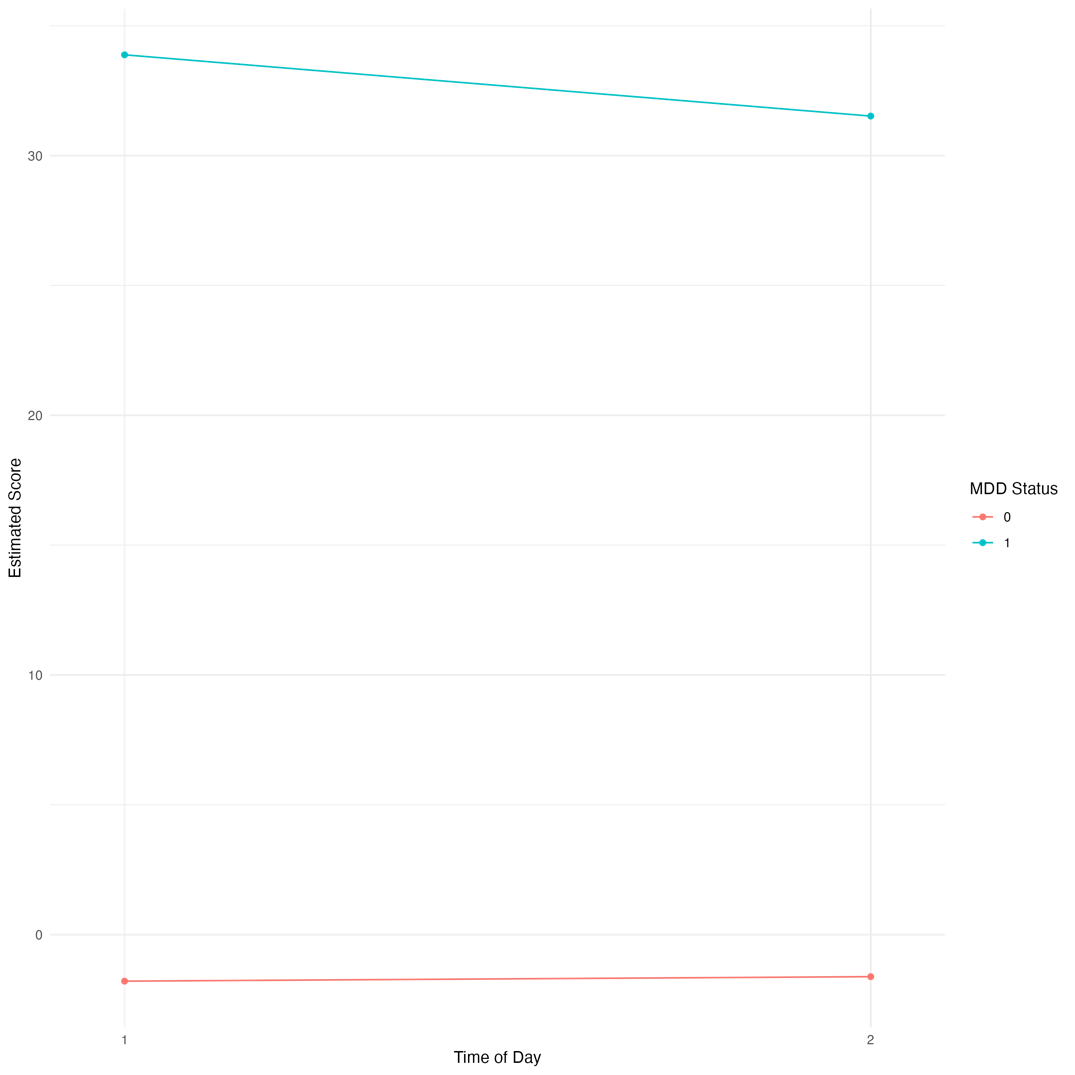

**Figure S2.** Interaction of time of day with MDD Status on Summed EMA Score. Data shown are estimated marginal means of summed EMA score from a mixed-effects regression model including random effects.

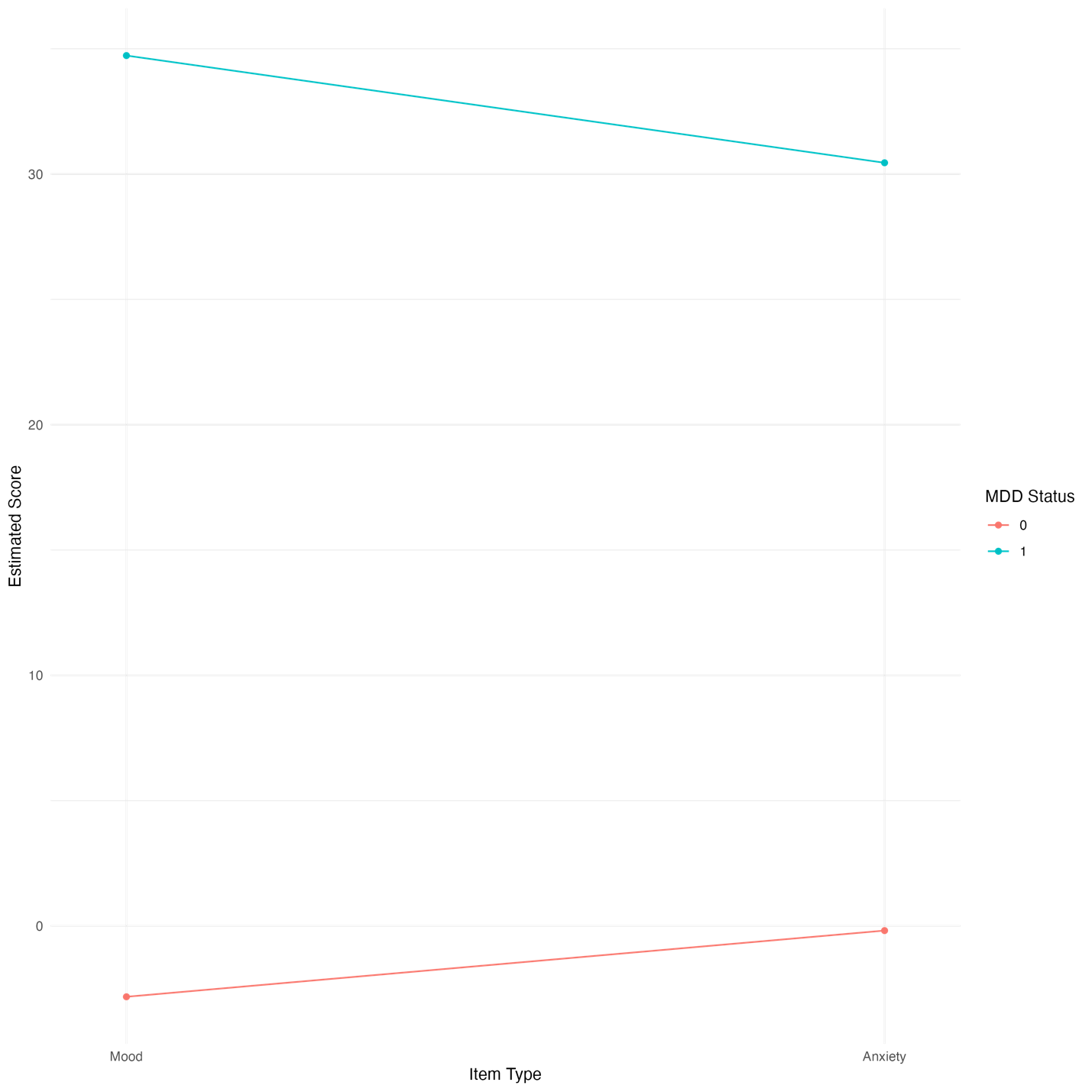

**Figure S3.** Interaction of item type with MDD Status on Summed EMA Score. Data shown are estimated marginal means of summed EMA score from a mixed-effects regression model including random effects.

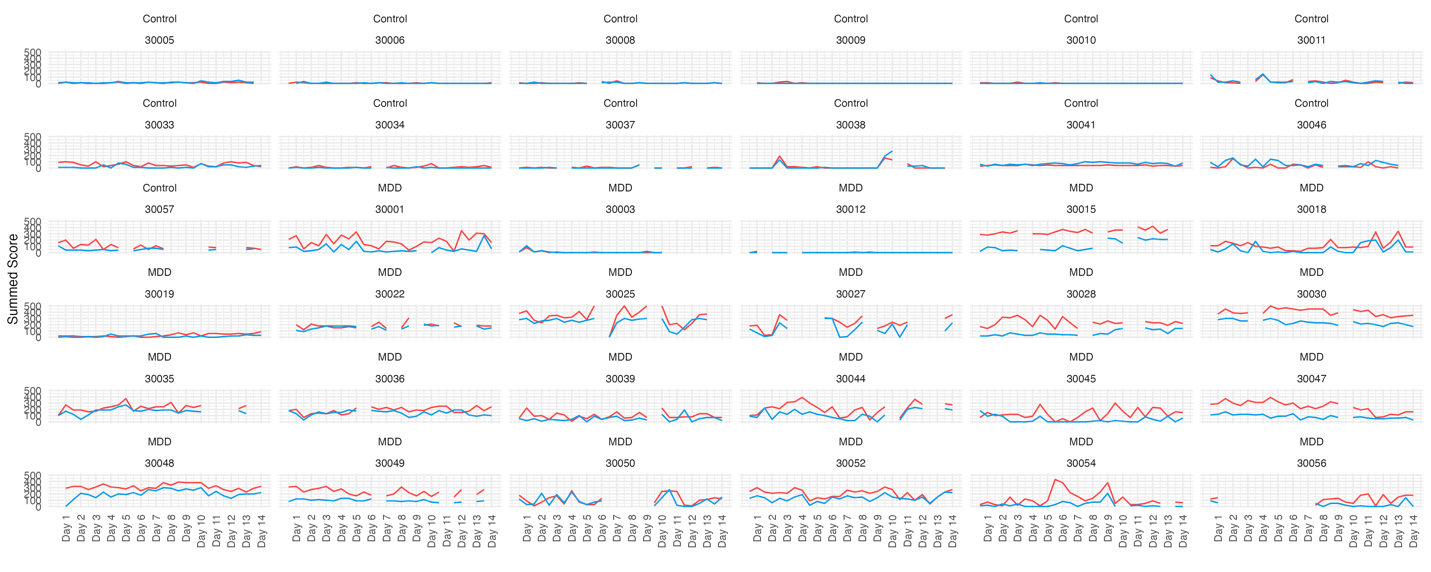

**Figure S4.** Facet plot of raw summed mood (red) and anxiety (blue) data over time in each participant.

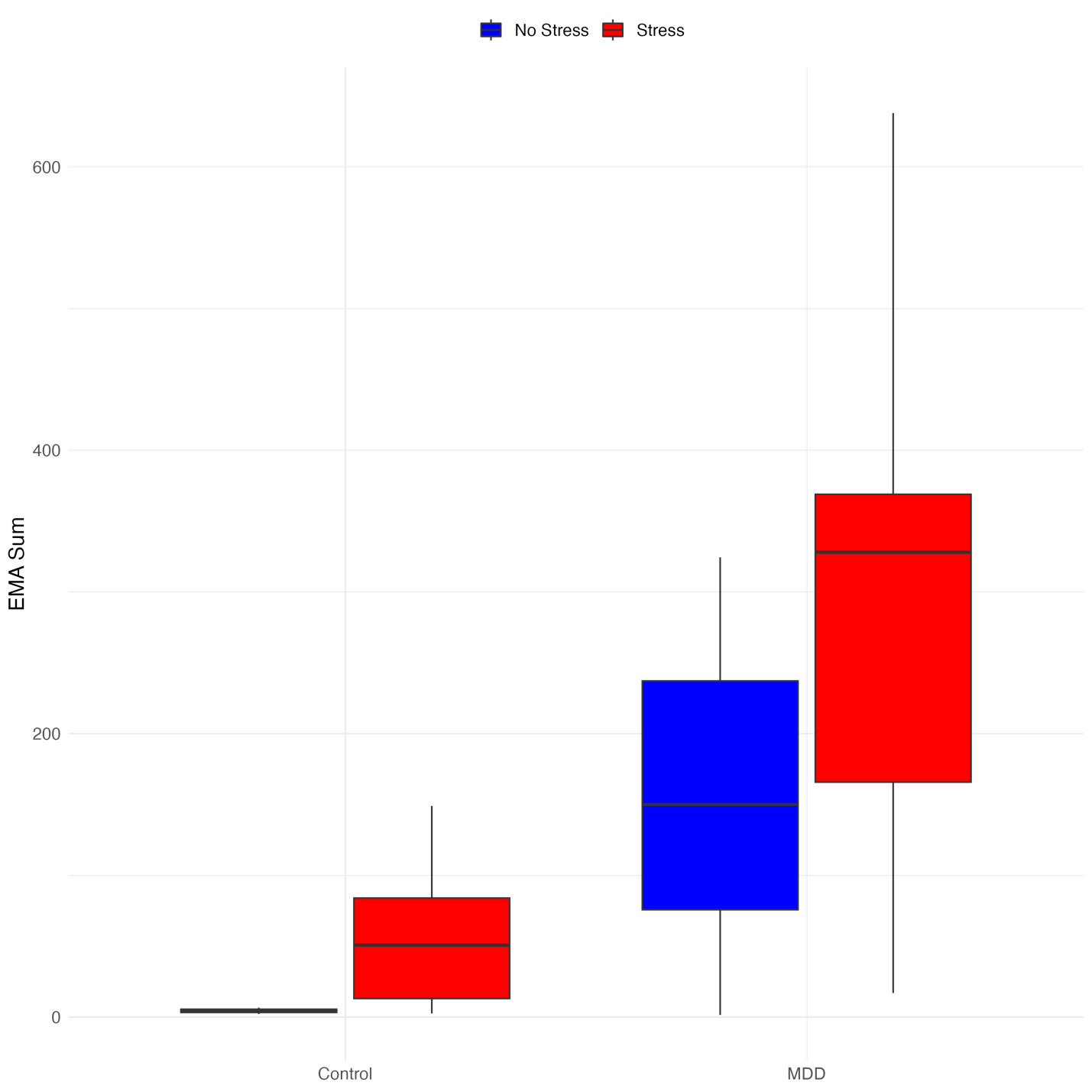

**Figure S5.** Boxplot of EMA sum score by group (MDD and control) and stress (no reported stress and reported stress).

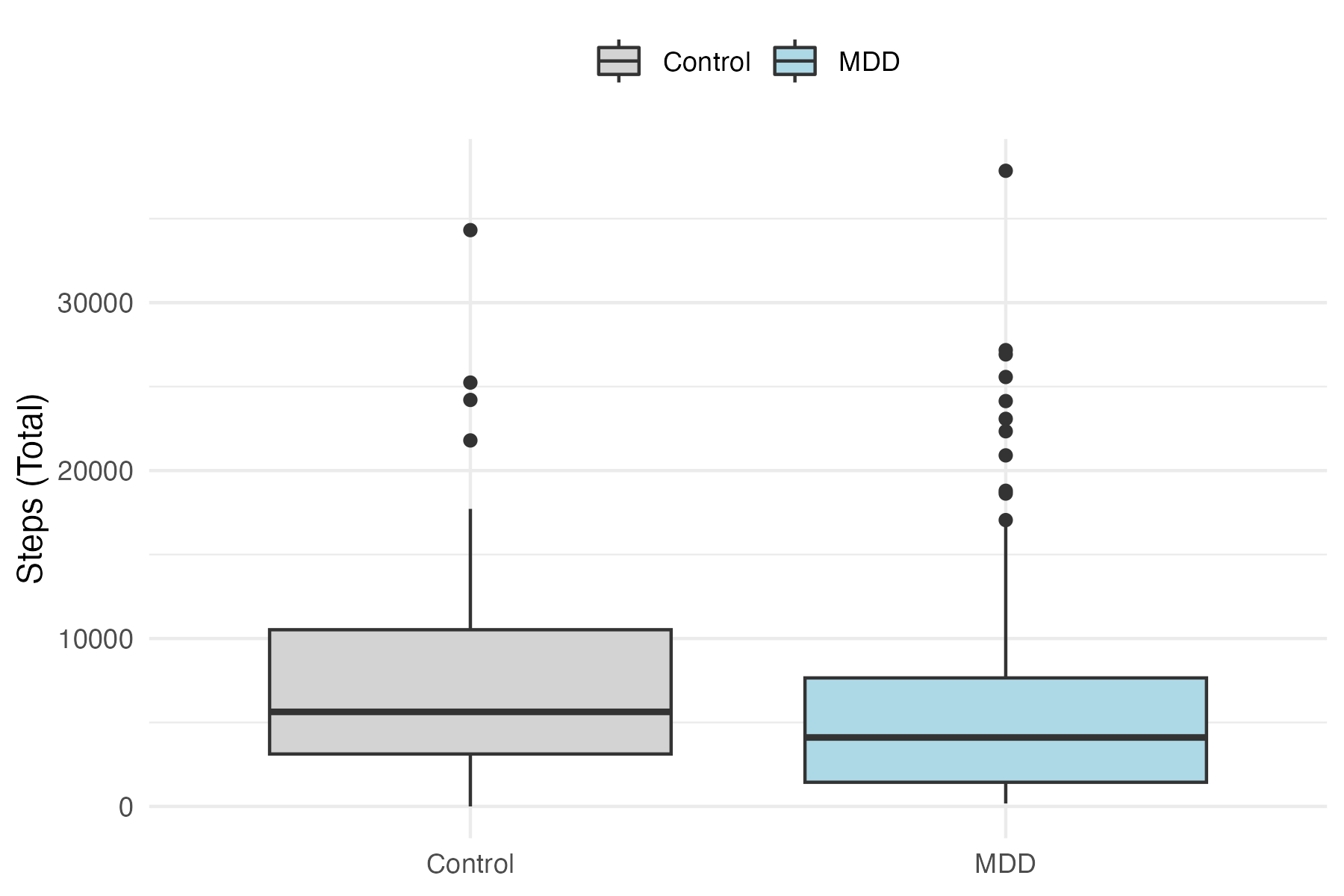

**Figure S6.** Boxplot of total steps taken during FitBit wearing by group (MDD and control).

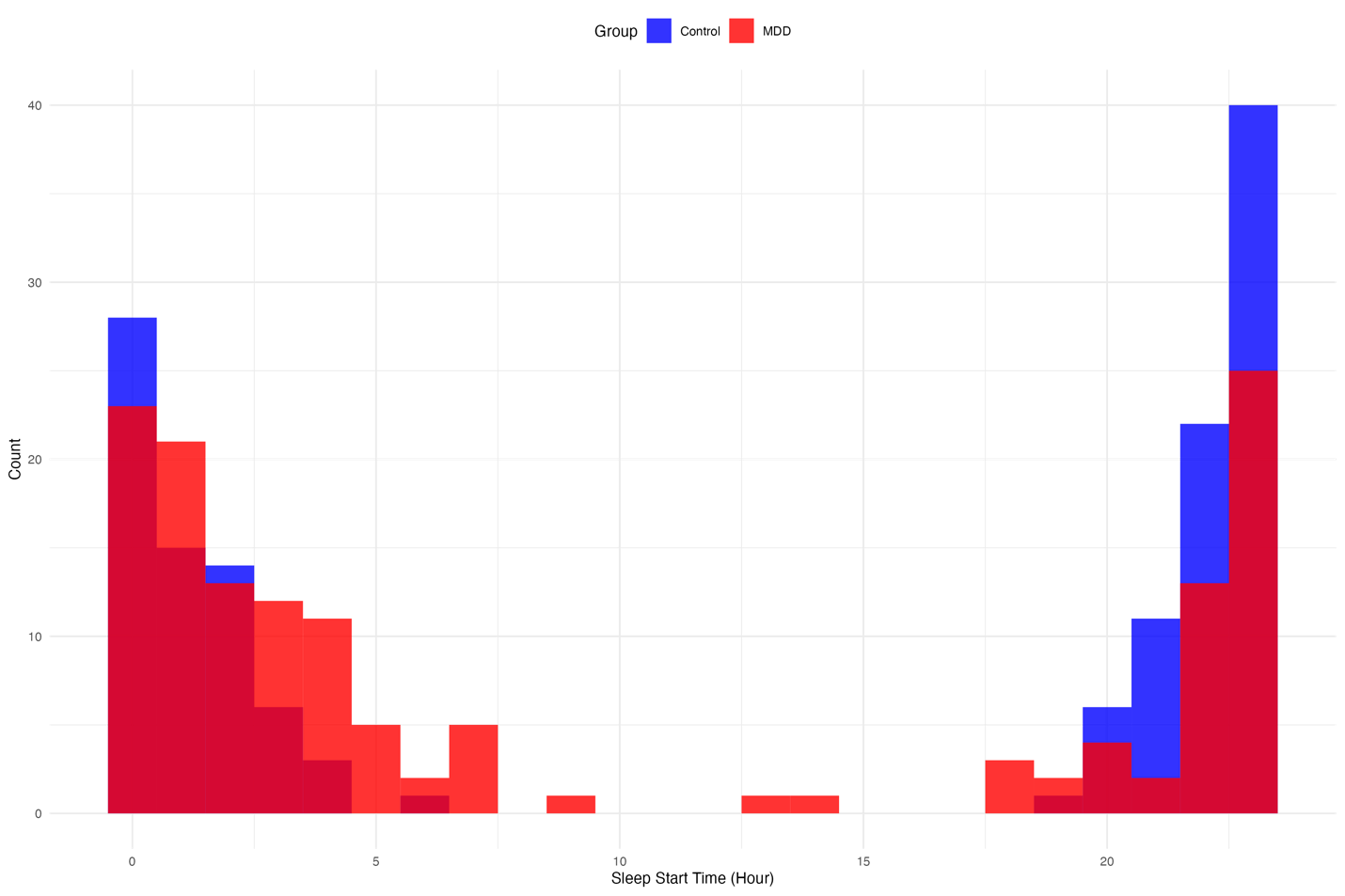

**Figure S7.** Histogram of sleep start times (rounded to closest hour) that were not naps (>=180 minutes) by group.

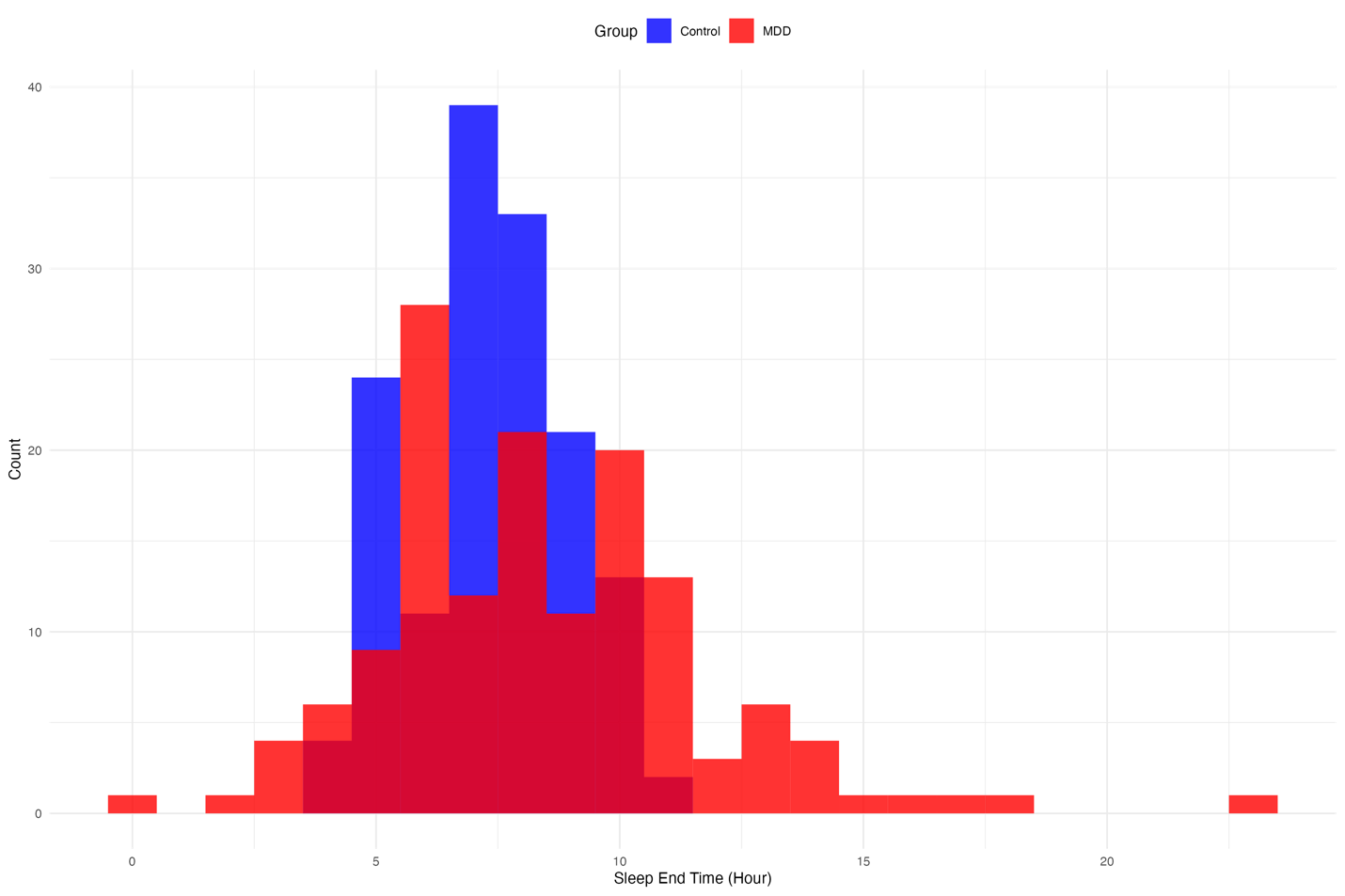

**Figure S8.** Histogram of sleep end times (rounded to closest hour) that were not naps (>=180 minutes) by group.

**Table S1.** Provenance of ecological momentary assessment (EMA) questions. All items originated from the PHQ-9 and GAD-7 but were edited to make them quicker to read and respond to, and suitable for twice daily use. Items were selected to cover the breadth of depressive and associate anxious symptomatology.

| EMA Item | Original Instrument | Wording in Original Instrument | Corresponds to Symptom |
| --- | --- | --- | --- |
| I am sad. | PHQ-9 | Over the past two weeks, how often have you felt down, depressed or hopeless. | Low Mood |
| I feel bothered by every little thing. | GAD-7 | Over the past two weeks, how often have you become easily annoyed or irritable. | Irritability |
| I have no interest in things I would usually enjoy (e.g., food, TV, games, spending time with friends/family). | PHQ-9 | Over the past two weeks, how often have you had little interest or pleasure in doing things. | Anhedonia |
| I do not have enough energy to get going. | PHQ-9 | Over the past two weeks, how often have you felt tired or had little energy. | Psychomotor - depression |
| I have no appetite or feel much hungrier than usual. | PHQ-9 | Over the past two weeks, how often have you had poor appetite or overeating. | Appetite |
| I feel physically tense and/or jittery. | GAD-7 | Over the past two weeks, how often have you been so restless that it is hard to sit still. | Psychomotor - anxiety |
| I am nervous, anxious or on edge. | GAD-7 | Over the past two weeks, how often have you felt nervous, anxious or on edge. | Anxiety |
| I can’t stop worrying | GAD-7 | Over the past two weeks, how often have you not been able to stop or control worrying. | Worry |

**Table S2.** Sleep features extracted from FitBit data per sleep log.

| **Features** |  | **Description** |
| --- | --- | --- |
| **Architecture** | TST | Total sleep time - minutes asleep |
|  | TIB | Time in bed – minutes in bed |
|  | TAB | Time awake in bed – minutes awake (sometimes referred to as WASO – wake after sleep onset) |
|  | Total Deep^*^ | Number of minutes in deep sleep |
|  | Total Light^*^ | Number of minutes in light sleep |
|  | Total REM^*^ | Number of minutes in REM sleep |
| **Quality** | Efficiency | Sleep efficiency (= minutes asleep / total minutes in bed) |
|  | Awake Five Mins | Number of awakenings >5 mins per sleep |
|  | Hypersomnia | Total sleep time > 10 hours |
|  | Naps | Number of sleeps lasting < 180 minutes |
| **Stability** | CoV TST | Coefficient of variation of total sleep time |
|  | SD Onset Time | Standard deviation of sleep onset time (time = minutes since 12 pm) |
|  | SD Offset Time | Standard deviation of sleep offset time (time = minutes since 12 pm) |

^*^ Excluding naps (<180 minute sleep length)

**Table S3.** Unconditioned model that includes EMA scores as DV and random effects.

| ***Predictors*** | ***Estimates*** | ***CI*** | ***p*** |
| --- | --- | --- | --- |
| (Intercept) | 22.18 | 13.50 – 30.86 | <0.001 |
| ***Random Effects*** |  | ***LRT*** | ***P*** |
| σ^2^ | 333.35 |  |  |
| τ_00_ _studyid:day:time:item_ | 54.36 | 100.79 | 1.02x10^-23^ |
| τ_00_ _studyid:day:time_ | 1.61 | 0.08 | 0.78 |
| τ_00_ _studyid:day_ | 62.12 | 102.23 | 4.94x10^-24^ |
| τ_00_ _studyid_ | 492.57 | 709.65 | 2.38x10^-156^ |
| ICC | 0.65 |  |  |
| N _studyid_ | 36 |  |  |
| N _day_ | 14 |  |  |
| N _time_ | 2 |  |  |
| N _item_ | 2 |  |  |
| Marginal R^2^ / Conditional R^2^ | 0.000 / 0.65 | |  |

**Table S4.** Frequency of life stressor reporting in MDD and control groups.

|  | **Control** | **MDD** |
| --- | --- | --- |
| I was not allowed to do something I wanted to do | 5 | 11 |
| I argued with a friend or family member | 11 | 21 |
| I got a bad grade in school. | 0 | 6 |
| I got disciplined or suspended from school | 0 | 0 |
| I did something that made me feel embarrassed | 10 | 23 |
| I was excluded from a group event | 0 | 5 |
| Someone in my family was arrested | 0 | 0 |
| Somebody in my family got a serious illness | 3 | 3 |
| Someone commented negatively on the way I look | 3 | 6 |
| My parents have been arguing a lot | 1 | 1 |
| Somebody teased or threatened me | 1 | 7 |
| I teased somebody else. | 1 | 8 |
| **Total** | **35** | **91** |

**Table S5.** Unique text responses after conversion to lowercase to item in the usability questionnaire “In which social settings did you use the app?”.

Response

out and about/errands

anywhere i was

around roommate

at dinner

at family's or girlfriend's house

at friend's house

at work/with friends

bar

bus

cafe

friend’s house

friend;s house

friends house

hanging out with family

home

house

in the car

literally anywhere.

mall

near friends/family

on break at work

on the bus

outside

restaurant

school

train

used it everywhere. had it over labor day weekend

usually kitchen

walking around

wherever notification went off

while traveling

work
